# The basic reproduction number of SARS-CoV-2: a scoping review of available evidence

**DOI:** 10.1101/2020.07.28.20163535

**Authors:** Ann Barber, John Griffin, Miriam Casey, Áine B. Collins, Elizabeth Ann Lane, Quirine Ten Bosch, Mart De Jong, David Mc Evoy, Andrew W. Byrne, Conor G. McAloon, Francis Butler, Kevin Hunt, Simon J. More

## Abstract

**Background:** The transmissibility of SARS-CoV-2 determines both the ability of the virus to invade a population and the strength of intervention that would be required to contain or eliminate the spread of infection. The basic reproduction number, R_0_, provides a quantitative measure of the transmission potential of a pathogen.

**Objective:** Conduct a scoping review of the available literature providing estimates of R_0_ for SARS-CoV-2, provide an overview of the drivers of variation in R_0_ estimates and the considerations taken in the calculation of the parameter.

**Design:** Scoping review of available literature between the 01 December 2019 and 07 May 2020.

**Data sources:** Both peer-reviewed and pre-print articles were searched for on PubMed, Google Scholar, MedRxiv and BioRxiv.

**Selection criteria:** Studies were selected for review if (i) the estimation of R_0_ for SARS-CoV-2 represented either the initial stages of the outbreak or the initial stages of the outbreak prior to the onset of widespread population restriction (“lockdown”), (ii) the exact dates of the study period were provided and (iii) the study provided primary estimates of R_0_.

**Results:** A total of 20 R_0_ for SARS-CoV-2 estimates were extracted from 15 studies. There was substantial variation in the estimates reported. Estimates derived from mathematical models fell within a wider range of 1.94-6.94 than statistical models which fell between the range of 2.2 to 4.4. Several studies made assumptions about the length of the infectious period which ranged from 5.8-20 days and the serial interval which ranged from 4.41-14 days. For a given set of parameters a longer duration of infectiousness or a longer serial interval equates to a higher R_0_. Several studies took measures to minimise bias in early case reporting, to account for the potential occurrence of super-spreading events, and to account for early sub-exponential epidemic growth.

**Conclusions:** The variation in reported estimates of R_0_ reflects the complex nature of the parameter itself, including the context (i.e. social/spatial structure), the methodology used to estimate the parameter, and model assumptions. R_0_ is a fundamental parameter in the study of infectious disease dynamics, however it provides limited practical applicability outside of the context in which it was estimated, and should be calculated and interpreted with this in mind.

**STRENGTHS AND LIMITATIONS OF THE SCOPING REVIEW:**

- This study provides an overview of basic reproduction number estimates for SARS-CoV-2 across a range of settings, a fundamental parameter in gauging the transmissibility of an emerging infectious disease.
- The key drivers of variation in R_0_ estimates and considerations in the calculation of the parameter highlighted across the reviewed studies are discussed.
- This evidence may be used to help inform modelling studies and intervention strategies.
- Given the need for rapid dissemination of information on a newly emerging infectious disease, several of the reviewed papers were in the pre-print phase yet to be peer-reviewed.

## INTRODUCTION

On 31 December 2019 a series of cases of pneumonia of unknown cause were notified to the World Health Organization (WHO). The *Coronaviridae* Study Group (CSG) of the International Committee of Taxonomy of Viruses designated this novel virus as SARS-CoV-2, the etiologic agent of coronavirus disease 2019 (COVID-19)[1]. While the origin of COVID-19 in humans has been attributed to spillover from an unknown wildlife source, human-human transmission has been responsible for the rapid spread of SARS-CoV-2 across the globe. On 11 March 2020, the WHO characterised COVID-19 as a pandemic[2].

An understanding of the transmissibility of a newly emerging infectious disease is required to determine the ability of a pathogen to spread and establish within a population along with the strength of mitigation required to contain or eliminate infection[3]. The basic reproduction number, R_0_, is an indicator of the transmissibility of an infectious agent, defined as the expected number of new infections that are generated, on average, by a single infected individual, over the course of its infectious period, in an otherwise uninfected population[4]. It is a threshold parameter: with a value above one, an infection can spread and persist within a population, below one infection cannot be sustained. R_0_ depends on the average contact rate between susceptible and infectious individuals (or infectious material of infectious individuals) per unit time, the (dimensionless) probability that the contact leads to infection and the average duration (time) of the infectious period[3]. The effective reproduction number, R, is a dynamic parameter that can chronicle the time-dependent (R_t_) variation in transmission as a result of, for example; the intrinsic decline in the proportion of susceptibles (e.g. infection and natural immunity) and, extrinsic effects of intervention on effective contact rates (e.g. social distancing), the probability that upon contact infection occurs (e.g. face masks) or the effective duration of infectiousness (e.g. self-isolation)[5]. It is an important parameter for monitoring the effectiveness of control measures and determining whether further measures are required. For the purpose of this review, we focus primarily on the basic reproduction number, R_0_.

The aim of this study was to conduct a scoping review of the available literature providing estimates on the basic reproduction number of SARS-CoV-2, to gain an understanding of the transmissibility of the virus and to aid the parameterisation of COVID-19 epidemiological models. R_0_ is a complex parameter of which the calculation and interpretation requires significant consideration. As such, we discuss and aim to disentangle several relevant concepts highlighted across the reviewed papers including the context, methodological differences, the relationship between R_0_ and the generation time/serial interval, the relationship between R_0_ and the infectious period, managing surveillance bias, individual variation in transmission and exponential epidemic growth.

## MATERIALS AND METHODS

### Search methodology, initial screening and categorisation

A survey of the literature between 01 December 2019 and 07 May 2020 for all countries was implemented, as part of a larger research project characterising key parameters of COVID-19[6–10]. The following search strategy was used. Publications listed in the electronic databases PubMed, Google Scholar, MedRxiv and BioRxiv were searched with the following keywords: (“Novel coronavirus” OR “SARS-CoV-2” OR “2019-nCoV” OR “COVID-19”) AND “reproduction number”. The dynamic curated PubMed database “LitCovid” was also monitored, in addition to national and international government reports. No restrictions on language or publication status were imposed so long as an English abstract was available. On the 23 July 2020, pre-print studies were checked for changes in publication status and any subsequent changes in estimates (Supplementary Table 1.).

Articles were evaluated for data relating to the aim of this review, and all relevant publications were considered for possible inclusion. Bibliographies within these publications were also searched for additional resources. This study is reported in compliance with the Preferred Reporting Items for Systematic Reviews and Meta-Analyses – Extension for Scoping Reviews (PRISMA-ScR) checklist[11].

### Study appraisal and selection for review

Studies were selected for review if they met the following criteria: (i) the estimation of R_0_ represented either the initial stages of the outbreak or the initial stages of the outbreak prior to the onset of widespread population restriction (“lockdown”), (ii) the exact dates of the study period were provided, and (iii) the study provided primary estimates of R_0_. A period prior to lockdown was selected as a compulsory requirement to ensure estimates provided were calculated under the realisation of an entirely susceptible population and any imposed disturbance on effective contact rates or infectious periods were minimised, as per the definition of R_0._ Study selection was undertaken by the first author. Parameter estimates for R_0_ and confidence intervals (where provided) were recorded and assessed. The study period, location of the study, methodology and model assumptions were also extracted.

## RESULTS

### Study appraisal and selection for review

There were 33 studies available for appraisal as of 07 May 2020 (Fig. 1).

**Figure 1.**
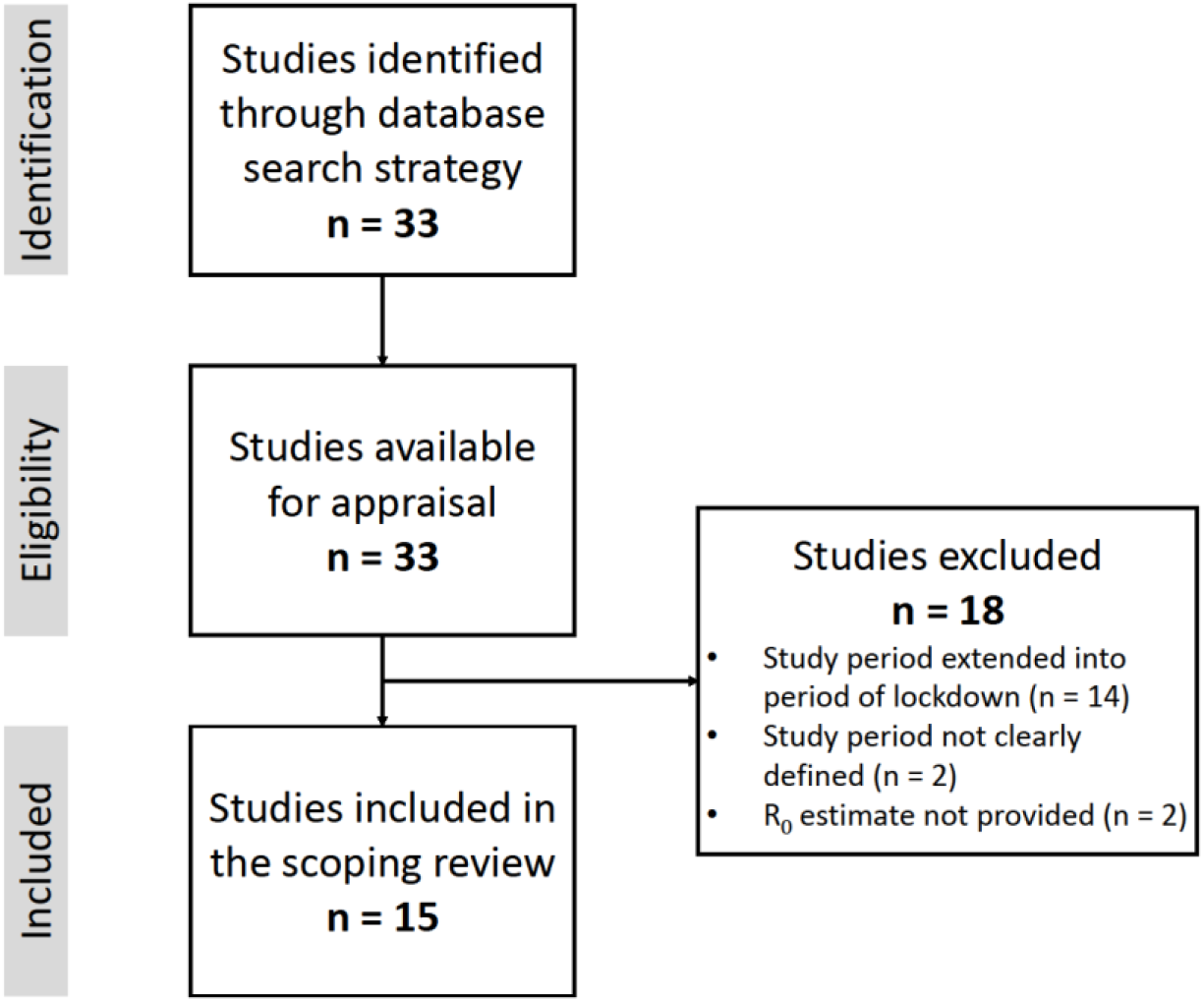
Flowchart for the studies included in the scoping review.

- 14 papers were removed as the study period extended into a period of lockdown[12–25].
- 1 paper was excluded as the study period was not clearly defined[26].
- 1 paper estimated R_0_ through phylogenetic analysis of 52 full genomes of viral strains sampled across different countries[27]. As this paper did not deal with a single population with defined study periods, it was not considered for review.
- 2 papers were excluded as they only considered the time-dependent reproduction number[28,29].

Following the removal of these papers, 15 papers were further evaluated, with estimates on the basic reproduction number provided in Table 1. Across these studies, estimates of R_0_ are provided for populations within China (n = 6 studies), Italy (n = 3 studies), Iran (n = 2 studies), the Republic of Korea (n = 1 study), France (n = 1 study), the United Kingdom (n = 1 study), Spain (n = 1 study) and onboard the Diamond Princess cruise ship (n = 1 study).

**Table 1.**
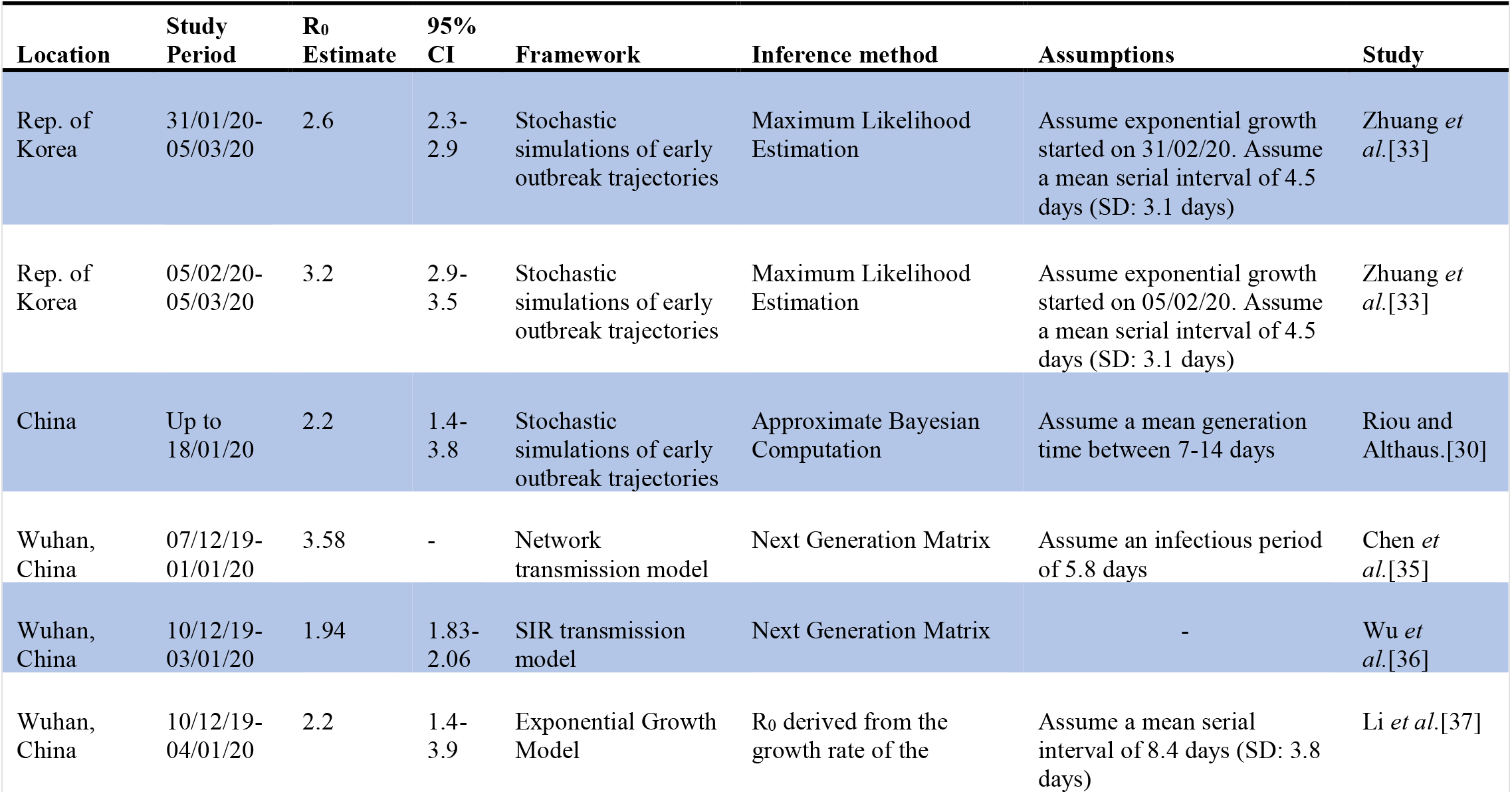
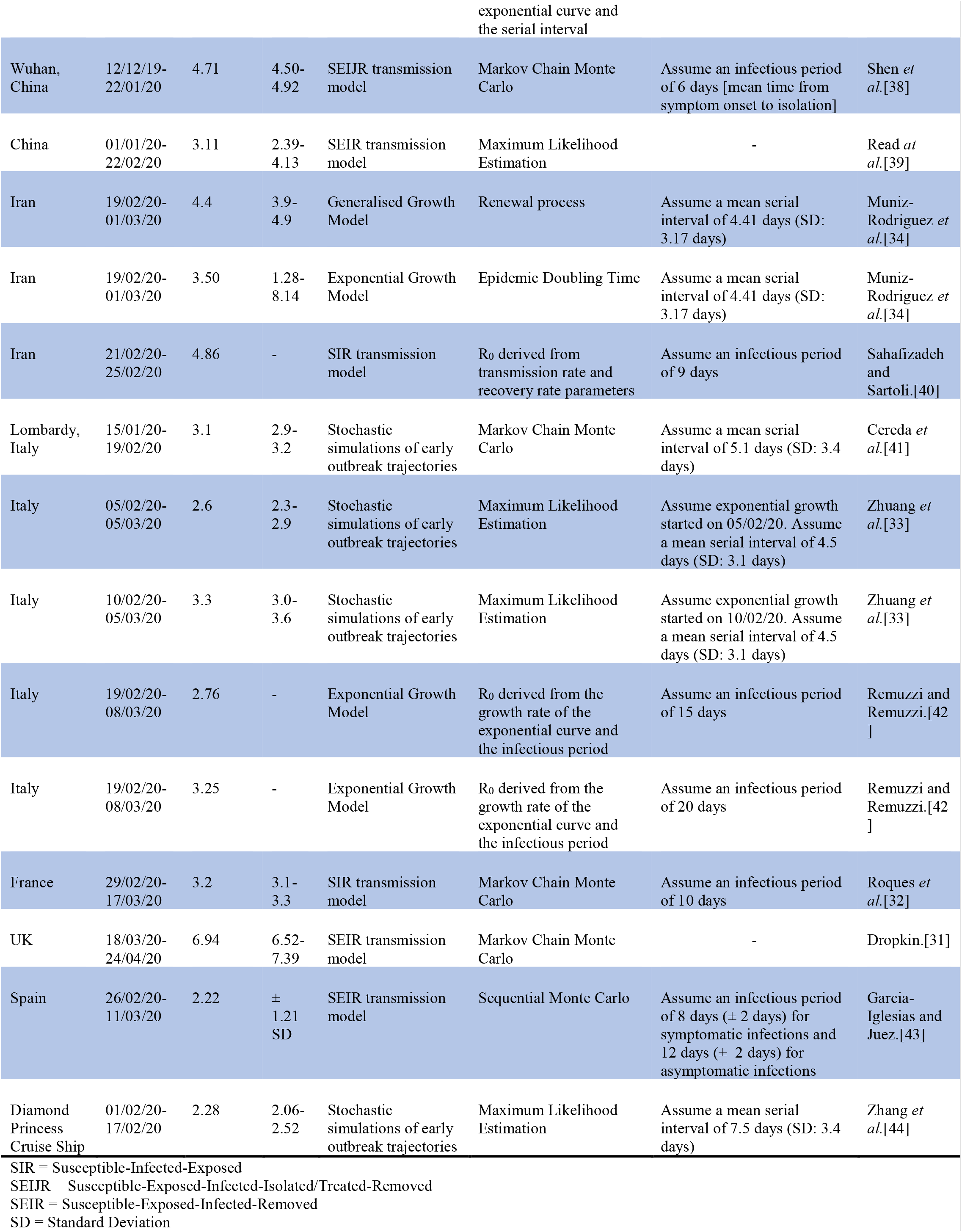
Estimates of the basic reproduction number, R_0_, provided by the 15 reviewed studies.

### Overall findings

From the 15 studies included, a total of 20 R_0_ estimates were reported, ranging from 1.94 to 6.94 (Table 1). R_0_ estimates varied both within countries, (for example, estimates in China, ranged between 1.94 and 4.71), and between countries (for example, an R_0_ of 2.2 was reported for Spain, 3.2 for France and 6.94 for the UK) (Fig. 2).

**Figure 2.**
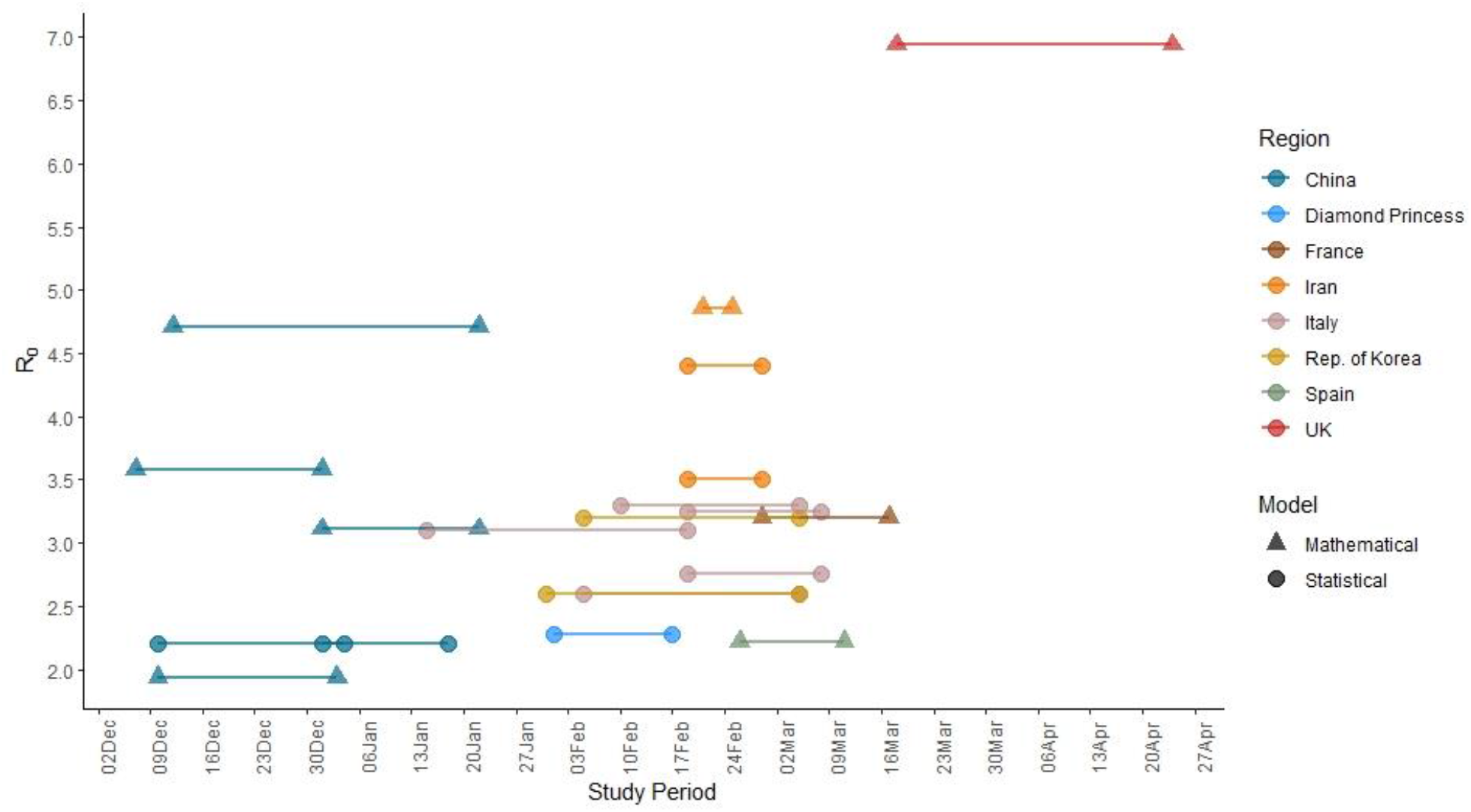
Relationship between start and end dates of the study period and estimated R_0_ across region and model.

Seven studies deployed statistical models and eight mathematical models (see Supplementary Table 2 for further methodological details). There were 2 types of statistical frameworks and 4 types of mathematical frameworks used across the studies. Overall, there were 10 types of inference methods used, 2 of which (Markov Chain Monte Carlo (MCMC) and Maximum Likelihood Estimation (MLE)) were used within both statistical and mathematical frameworks while 5 types of inference methods were used exclusively within statistical frameworks and three within mathematical frameworks.

Assumptions surrounding the duration of the infectious period and the serial interval were present across several of the studies. Assumptions about the length of the infectious period ranged between 5.8 to 20 days and assumptions on the serial interval ranged between 4.4 to 14 days. Three studies applied methods to control for surveillance bias in case reporting[30–32], the potential occurrence of individual variation in transmission was included in two studies[30,33], and one study accounted for sub-exponential growth in the early stages of the outbreak[34].

## DISCUSSION

In this review we found a total of 20 reported R_0_ estimates from 15 studies which ranged from 1.94 to 6.94. There was substantial variation in estimates both within and between countries. We found 12 values of R_0_ were estimated within statistical frameworks and 8 values were estimated within mathematical frameworks. The duration of the infectious period and the serial interval were common assumptions ranging in value across several studies. We identified three key potential drivers of variation in reported estimates of R_0_ including: i) the complex nature of R_0_ itself, in that it is context specific, ii) methodological differences and iii) the model assumptions (including the duration of the infectious period and serial interval). Further we discuss how studies sought to minimise surveillance bias, the importance of individual variation in transmission, and the occurrence of exponential growth at the early stages of an outbreak.

### Drivers of variation

#### i) Context

A key component of R_0_ is the effective contact rate (a function of the average contact rate and probability that contact leads to infection), which can vary over space or time. Traditional epidemiological models assume homogeneous mixing of individuals, where equal contact rates are assumed between individuals within a population. Applying this simplifying assumption allows for more tractable modelling and analysis but in reality heterogeneities in contact patterns are introduced by, for example; population density and social organisation[45]. In density dependent systems, transmission increases linearly with population density, and a threshold density is required for infection to persist[46]. For example; density dependent infections are more likely to propagate and persist in urban settings compared to rural. Social structure also plays an important role in infection transmission, where infectious individuals are more likely to infect members of the same social group[47]. R_0_ is therefore considered a function of the factors capable of influencing contact rates along with the biological components of the pathogen (e.g. the probability that contact leads to infection and the length of the infectious period)[48]. R_0_ provides limited practical applicability outside of the context in which it was estimated; therefore, R_0_ for one geographical/social setting may not be applicable to that of another[49]. Even within the same region variation in estimates occur. To illustrate, 4 estimates were reported for Wuhan, which ranged from 1.94-4.71.

#### ii) Methodological differences

Statistical and mathematical models in epidemiology play a fundamental role in providing an understanding of transmission dynamics and evaluating the effectiveness of control strategies. Statistical methods to estimate R_0_ which rely on incidence data and are generally in the form of either descriptive non-dynamical models or stochastic models. At the early phase of an epidemic, the number of new infections per unit time increases exponentially and R_0_ can be estimated in descriptive non-dynamical models, for example exponential growth models, from the empirically observed growth rate *p* of the epidemic curve[50,51]. In contrast, stochastic models, for example branching process models, simulate early outbreak trajectories based on the generation concept whereby each infected individual *i* is associated with a generation time *T*_*G*_. At the end of *T*_*G*_, a random number *N*_*i*_ of new infections (‘offspring’) have been generated, where R_0_ is the offspring mean[52–54]. Furthermore, where data on chains of transmission (i.e. who infected whom) are available, the number of secondary infections generated by each infected individual can be simply counted. In the current study values of R_0_ estimated by means of statistical methods fell within the range of 2.2-4.4.

Fitting mechanistic models to epidemic data can provide useful insights about transmission dynamics. These models are capable of describing the unobservable mechanism of transmission between individuals whereby dynamics at the population level are composed of the sum of individual-level processes of which the observed data are generated. Furthermore, they provide a suitable framework to study the effects of control measures on the spread of infection[52,55,56]. R_0_ can be calculated directly from model parameters or coupled with statistical techniques[57]. In the current study, R_0_ values estimated by mathematical models fell within the range of 1.94 to 6.94, a substantially wider range compared to that of estimates generated within statistical frameworks. Transmission models range in complexity depending on the types of questions addressed, subject to trade-off between structuring the model to conform with biological reality and the ability to sensibly parameterise and generalise the system[58]. As such the wider variation across these models may be attributed to model structure or assumptions, along with the context in which R_0_ was estimated.

Several studies deployed a set of ordinary differential equations to model disease spread[31,32,35,36,38–40,43]. Parsimonious models may be robust to difficulties in model parameterisation associated with limited data availability[32,40]. However, as a consequence, they lack the ability to capture more realistic dynamics such as pre- and asymptomatic transmission, susceptibility to different levels of clinical severity in infection, or age-related infection dynamics. In contrast, a reservoir-people transmission network model simulating the spread of infection captured spill-over from a wildlife reservoir and human-human transmission as well as incorporating variation in transmission between symptomatic and asymptomatic infected individuals[35]. Although this model describes the detailed process of infection spread associated with SARS-COV-2, model parameterisation is heavily dependent on limited data sourced from the literature, unreliable mobility data, and assumptions surrounding the proportion and relative transmissibility of asymptomatic infection. Complex models may provide a good fit to the data, however, any relationship with the underlying process may not necessarily be interpretable. Further, complex models may overfit the data and thus; generalise poorly. In general, parsimonious models are preferred to complex ones and models should not contain more than the minimum necessary assumptions[59]. While model structure may, in part, account for variation in R_0_ estimates, even within models with similar structure variation in estimates occur, reflecting differences in the context or model assumptions.

#### iii) Model assumptions

##### On the relationship between R_0_ and the duration of the infectious period

The infectious period, the time between the end of the pre-infectious period to when an individual can no longer pass on infection, is a key component determining R_0_. It is a clinical characteristic which does not necessarily differ across settings. With a newly emerging infectious disease it is often difficult to quantify the duration of the infectious period where relatively little is known in relation to the occurrence or relative occurrence of events such as asymptomatic, pre-symptomatic and symptomatic infectiousness. As such, several studies made assumptions about the duration of the infectious period which ranged from 5.8 to 20 days. One study derived R_0_ from the growth rate of the epidemic curve and the duration of the infectious period which was assumed to be either 15 or 20 days[42]. For a given growth rate, an infectious period of 20 days resulted in a higher R_0_ (R_0_ = 3.25) than a shorter infectious period of 15 days (R_0_ = 2.76). A longer infectious period relates to a longer time for potential transmission opportunities to occur. The inverse of the average duration of infectiousness is the recovery rate (that is, how quickly an individual exists a state of infectiousness) and so a shorter infectious period will result in individuals exiting the infectious state at a faster rate than that of a longer infectious period. Therefore, for a given set of parameters, a longer duration of infectiousness equates to a higher expected R_0_ than that associated with a shorter infectious period.

##### On the relationship between R_0_ and the generation time/serial interval

Several studies rely on the distribution of the generation time or its proxy the serial interval (time between the infection events or symptom onset of primary and secondary cases, respectively) in the estimation of R_0_[30,34,37,41,44]. The serial interval is a context-specific parameter that is sensitive to factors such as contact patterns. For example, if transmission following symptom onset is reduced due to isolation of symptomatic individuals, there may be relatively more pre-symptomatic transmission events and shorter serial intervals[9]. Ideally, the serial interval should be estimated from the corresponding population of interest. However, in reality this may not always be feasible, particularly at the early stages of an outbreak where data may be limited in availability. When there is a scarcity of data, studies often make assumptions about the distribution of the serial interval. This included assuming a serial interval distribution based on previous reports[34], and exploring a range of parameter combinations that include the distribution of the serial interval[30]. Two studies directly estimated the distribution of the serial interval by fitting a gamma distribution to data from the population under study[37,41]. One study investigated the sensitivity of the time-dependent reproduction number, R_t_, to the serial interval and showing that although R_t_ followed the same trend across values of the serial interval, a longer serial interval resulted in a higher R_t_ estimate[41]. For a given rate of epidemic growth, a higher R will require fewer (so longer) generations of transmission chains to realise the same population of infection spread that would occur with a lower R (i.e. a shorter generation interval equates to faster transmission at the individual level)[60].

### Minimising surveillance bias

There are several challenges associated with incidence data at the early epidemic stages of a newly emerging infectious disease. Case reporting is often unstable during this period due to lack of knowledge or awareness of, or confusion with, case definitions, which can limit the ability of health officials to identify infected individuals. Further, during the early stages of an epidemic, diagnostic facilities are often lacking in quality and/or quantity. These factors may contribute to unreliable or incomplete data limiting the ability to accurately characterise important parameters, such as R_0_, with high levels of precision early in an outbreak. If the rate of reporting remains stable over time, estimates of the reproduction number would be unaffected by underreporting[61]. However, as awareness increases, case definitions change and diagnostic facilities improve, variations in reporting over time may lead to erroneous interpretations of the reproduction number. Several of the reviewed studies took measures to minimise surveillance bias, including smoothing case and death data with a moving average over 5 days[32], gaining an indirect estimate of epidemic size through cases identified outside of the study population[30], and relying on hospital death data[31], which may be more stable than case reporting. These methods provide alternatives to relying on unreliable or unstable surveillance data at the early stages of an outbreak.

### Individual variation in transmission

Superspreading events (SSEs) describe situations where a small proportion of infectious individuals account for more than the expected number of transmission events. As R_0_ describes a population-average of the number of secondary cases generated by a primary case, individual-level variation in transmission is not expressed. Greater levels of individual heterogeneity in transmission lead to a less frequent occurrence of outbreaks, however outbreaks that do occur, have a tendency to occur explosively[62,63]. Several cases of COVID-19 SSEs have been identified[64–66]. While SSEs often remain rare events, they can have important implications for the epidemic trajectory and control efforts[30]. Following the approach described by Lloyd-Smith *et al*., (2005)[62], in the estimation of R_0_, two studies incorporated a dispersion parameter, *k*, that measures the likelihood of occurrence of SSEs[30,33]. The lower the value of *k*, the greater the level of individual heterogeneity in transmission. Both studies indicated that homogenous patterns of transmission events were more likely, although suggested that the potential for SSEs to occur should not be discounted[30]. These results are inconsistent with a study modelling overdispersion in transmission of COVID-19, which suggested that approximately 10% of infectious individuals were responsible for 80% of secondary transmissions where R_0_ values ranged between 2 and 3[67]. Overdispersed transmission (high individual-level variation) has important implications for control measures, for example if transmission is highly overdispersed control efforts would be most effective by targeting situations where SSEs are likely to occur.

### Exponential growth

At the early stages of an epidemic, while the depletion of susceptibles is negligible the number of new cases per unit time grows exponentially. The growth rate of the exponential curve measures how quickly infection spreads and can be used to estimate R_0_. However, if the initial growth phase is slower than exponential (i.e. sub-exponential growth), R_0_ estimates from exponential growth models may be inflated. The generalised growth model is capable of explicitly accounting for a sub-exponential growth phase[68,69]. One study applied this framework incorporating a scaling factor (the deceleration of growth parameter[68]) to account for the sub-exponential growth phase[34]. Although the effect of different scaling factor values on R_0_ was not explored, it is expected that the further that epidemic growth is from exponential, the lower R_0_ would be[68]. Consideration should therefore be given as to whether the early stage of the epidemic is truly growing exponentially when applying exponential growth models to estimate R_0_.

## CONCLUSION

We found 20 reported R_0_ estimates which ranged from 1.94 to 6.95. We identified several key drivers of variation in estimates including the context (i.e. social/geographic structure), methodology (i.e. mathematical vs statistical frameworks and model structure) and model assumptions (i.e. duration of the infectious period and serial interval). Notably, there was a wider variation in estimates generated within a mathematical framework (1.94-6.94) compared to a statistical framework, which fell within a narrower range of 2.2 to 4.4. However, variation across estimates is unlikely to be a result of a single driver in variation rather estimates differ across drivers or combinations of drivers. Here we have highlighted key potential sources of variation in R_0_ and attempted to disentangle the effects these drivers have on the estimates generated. Further, we identified several key considerations accounted for, including: approaches to minimise bias introduced by unstable case reporting, the importance of individual variation in transmission and the early epidemic characterisation of exponential or sub-exponential growth. R_0_ is a fundamental parameter in the study of infectious disease dynamics however the calculation and interpretation is not a straightforward exercise and careful consideration is required for both. Although this review details the some of these considerations, we have by no means exhausted all potential drivers of variation and we have not discussed all of the complexities associated with R_0_. However, we have discussed the key considerations that occurred within the reviewed studies in the context of COVID-19 and the theory is widely applicable across the board of infectious diseases which may help inform future studies attempting to calculate R_0_.

## Data Availability

N/A

## FOOTNOTES

### Author contributions

AB conducted the eligibility screening of shortlisted studies, extracted the data and reviewed the parameter estimates with input from all authors; ÁC, KH and FB conducted the initial literature searches; AB completed the initial drafts of the manuscript; All authors read earlier manuscript versions and approved the final manuscript.

### Patient and public involvement statement

It was not appropriate or possible to involve patients or the public in the design, or conduct, or reporting, or dissemination plans of our research.

## SUPPLEMENTARY MATERIAL

**Supplementary Table 1:** Initial and updated estimates of the basic reproduction number, R_0_. All studies initially reviewed (extracted up to 07 May 2020) in the pre-print phase were revisited on 23 July 2020 to check for updates in publication status and/or changes in reported estimates. 5 pre-print studies remain in the pre-print phase without changes to estimates of R_0_.(1–5) 1 pre-print study was subsequently peer-reviewed with no changes to the initial estimate.(6) 2 pre-print studies were subsequently peer-reviewed with changes to the initial reported estimates – in both cases there were changes to the study period and thus, data informing the analyses.(7,8) One study initially provided estimates for both the Republic of Korea and France, the peer-reviewed version did not contain the Rep. of Korea analysis and there was a change in the estimate reported for France (initially assumed an infectious period of 20 days, subsequently reduced to 10 days).(9)

| **Supplementary Table 1. Initial and updated estimates of the basic reproduction number, $R_0$.** |  |  |  |  |  |  |  |
| --- | --- | --- | --- | --- | --- | --- | --- |
| Paper | Initial $R_0$ Estimate | Initial 95% CI | Initial Status | Updated $R_0$ Estimate | Updated 95% CI | Updated Status | Comments |
| Garcia-Iglesias and Juez.(1) | 2.22 | ± 1.21 SD | Pre-print | 2.22 | ± 1.21 SD | Pre-print | No changes to initial estimates |
| Read *et al.*(2) | 3.11 | 2.39-4.13 | Pre-print | 3.11 | 2.39-4.13 | Pre-print | No changes to initial estimates |
| Shen *et al.*(3) | 4.71 | 4.50-4.92 | Pre-print | 4.71 | 4.50-4.92 | Pre-print | No changes to initial estimates |
| Cereda *et al.*(4) | 3.1 | 2.9-3.2 | Pre-print | 3.1 | 2.9-3.2 | Pre-print | No changes to initial estimates |
| Sahafizadeh and Sartoli.(5) | 4.86 | - | Pre-print | 4.86 | - | Pre-print | No changes to initial estimates |
| Zhuang *et al.*(6) | 2.6 | 2.3-2.9 | Pre-print | 2.6 | 2.3-2.9 | Peer-reviewed | No changes to initial estimates |
|  | 3.2 | 2.9-3.5 |  | 3.2 | 2.9-3.5 |  |  |
|  | 2.6 | 2.3-2.9 |  | 2.6 | 2.3-2.9 |  |  |
|  | 3.3 | 3.0-3.6 |  | 3.3 | 3.0-3.6 |  |  |
| Muniz-Rodriguez *et al.*(8) | 3.6 | 3.2-4.2 | Pre-print | 4.4 | 3.9-4.9 | Peer-reviewed | Data updated from 19 Feb - 29 Feb to 19 Feb - 01 Mar |
|  | 3.58 | 1.29-8.46 |  | 3.5 | 1.28-8.14 |  |  |
| Dropkin.(7) | 5.81 | 5.08-6.98 | Pre-print | 6.94 | 6.52-7.39 | Peer-reviewed | Data updated from 28 Feb - 23 Mar to 18 Mar - 24 Apr |
| Roques *et al.*(9) | 2.6 | - | Pre-print | - | - | - | Korean analysis excluded. |
|  | 4.8 | - | Pre-print | 3.2 | 3.1-3.3 | Peer-reviewed | Initial estimate based on infectious period of 20 days, current estimate based on infectious period of 10 days. |

**Supplementary Table 2:**
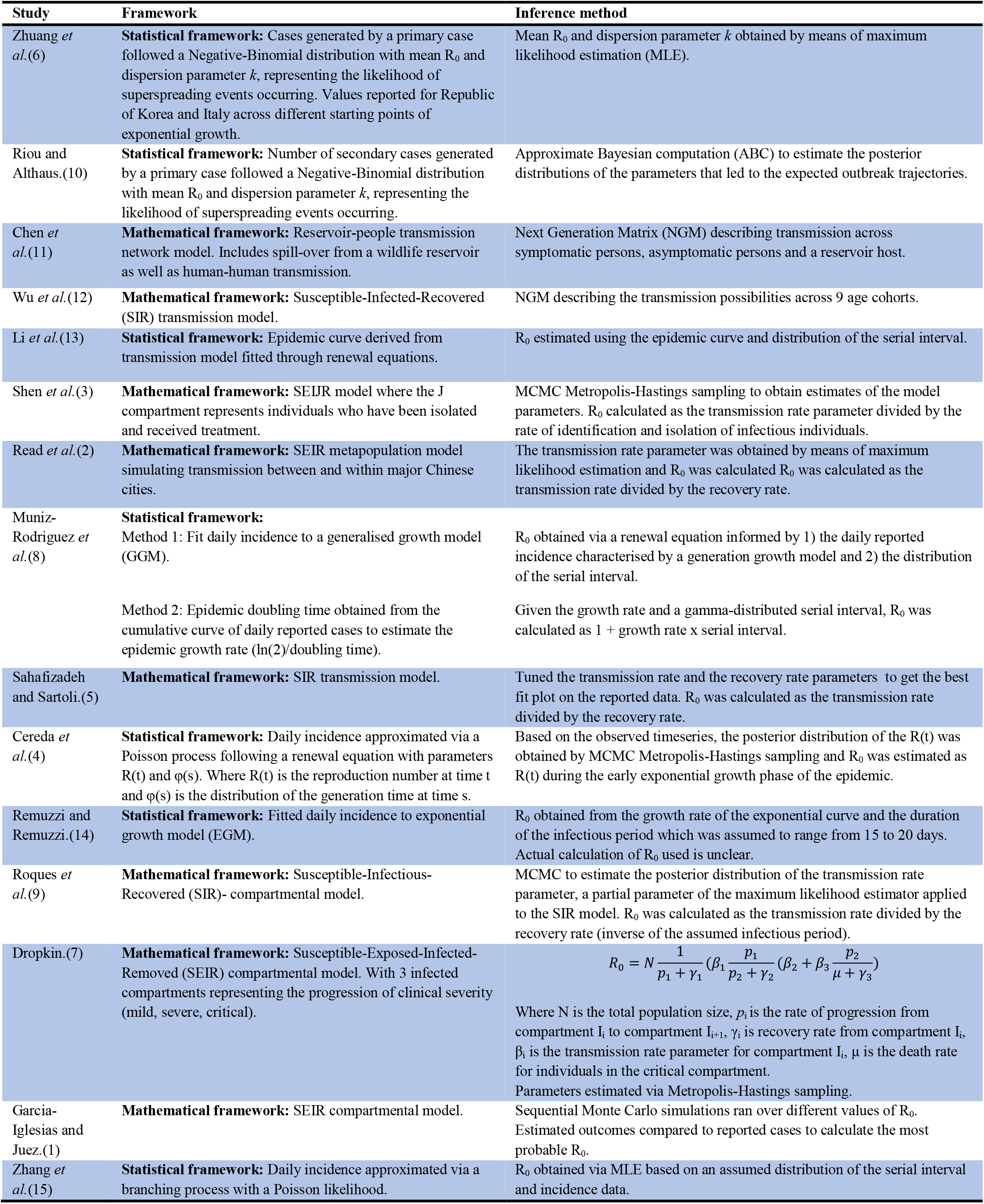
Summary of the framework and inference method adopted across the 15 reviewed studies. Studies were categorised into 2 frameworks based on the modelling approach: 1) mathematical models and 2) statistical models. Mathematical models were either in the form of compartmental epidemic models (e.g. Susceptible-Infectious-Recovered models) or network models (e.g. Reservoir-Human and Human-Human models). Statistical frameworks included those of epidemic growth models (e.g. exponential/generalised growth models) and stochastic simulations of early outbreak trajectories (e.g. branching process models). The inference method refers to the method of calculation of R_0_. Several approaches were used to calculate R_0_, for example; directly obtaining the parameter from models (e.g. Maximum Likelihood Estimation (MLE) of the mean (R_0_) of the offspring distribution) or deriving R_0_ from model parameters (e.g. the transmission rate parameter divided by the recovery rate parameter or 1 + growth rate X serial interval).

